# Early Safety Indicators of COVID-19 Convalescent Plasma in 5,000 Patients

**DOI:** 10.1101/2020.05.12.20099879

**Authors:** Michael J. Joyner, R. Scott Wright, DeLisa Fairweather, Jonathon W. Senefeld, Katelyn A. Bruno, Stephen A. Klassen, Rickey E. Carter, Allan M. Klompas, Chad C. Wiggins, John R.A. Shepherd, Robert F. Rea, Emily R. Whelan, Andrew J. Clayburn, Matthew R. Spiegel, Patrick W. Johnson, Elizabeth R. Lesser, Sarah E. Baker, Kathryn F. Larson, Juan G. Ripoll, Kylie J. Andersen, David O. Hodge, Katie L. Kunze, Matthew R. Buras, Matthew N.P. Vogt, Vitaly Herasevich, Joshua J. Dennis, Riley J. Regimbal, Philippe R. Bauer, Janis E. Blair, Camille M. Van Buskirk, Jeffrey L. Winters, James R. Stubbs, Nigel S. Paneth, Arturo Casadevall

## Abstract

**Background:** Convalescent plasma is the only antibody based therapy currently available for COVID-19 patients. It has robust historical precedence and sound biological plausibility. Although promising, convalescent plasma has not yet been shown to be safe as a treatment for COVID-19.

**Methods:** Thus, we analyzed key safety metrics after transfusion of ABO-compatible human COVID-19 convalescent plasma in 5,000 hospitalized adults with severe or life-threatening COVID-19, with 66% in the intensive care unit, as part of the US FDA Expanded Access Program for COVID-19 convalescent plasma.

**Results:** The incidence of all serious adverse events (SAEs) in the first four hours after transfusion was <1%, including mortality rate (0.3%). Of the 36 reported SAEs, there were 25 reported incidences of *related* SAEs, including mortality (*n*=4), transfusion-associated circulatory overload (TACO; *n*=7), transfusion-related acute lung injury (TRALI; *n*=11), and severe allergic transfusion reactions (*n*=3). However, only 2 (of 36) SAEs were judged as definitely related to the convalescent plasma transfusion by the treating physician. The seven-day mortality rate was 14.9%.

**Conclusion:** Given the deadly nature of COVID-19 and the large population of critically-ill patients included in these analyses, the mortality rate does not appear excessive. These early indicators suggest that transfusion of convalescent plasma is safe in hospitalized patients with COVID-19.

**Brief Summary:** After transfusion of COVID-19 convalescent plasma in 5,000 patients, the incidence of serious adverse events was <1% and the seven-day incidence of mortality was 14.9%.

## Introduction

The number of confirmed cases of coronavirus disease 2019 (COVID-19) and the number of deaths attributed to COVID-19 in the US exceed that of any other country in the world (1). The overall case fatality rate for diagnosed COVID-19, appears to be about 4% (2), and reports from Wuhan suggest case-fatality rates of 14% among hospitalized patients (3), and 57% among intensive care unit (ICU) admissions on ventilators or requiring a fraction of inspired oxygen > 60% (4). The reported fatality rate in the United States ranged from 21% in New York City hospitals (5) to 50% reported in an early case series from the Seattle area (6). In response to the COVID-19 outbreak in the US and reported case-fatality rates, the US Food and Drug Administration (FDA) in collaboration with the Mayo Clinic and national blood banking community developed a national Expanded Access Program (EAP) to collect and distribute convalescent plasma donated by individuals that have recovered from COVID-19. There is historical precedent to anticipate that human convalescent plasma is a viable option for mitigation and treatment of COVID-19 (7, 8). Human convalescent plasma uses antibodies harvested from recently-infected and currently-recovered COVID-19 patients to treat currently-infected COVID-19 patients. This approach is referred to as passive antibody therapy. As recently summarized (7), convalescent plasma represents a promising treatment strategy with strong historical precedence, biological plausibility, and limited barriers for rapid development and deployment of this investigational therapy.

Passive antibody therapy was first described in the 1890s as the only means of treating certain infectious diseases prior to the development of antimicrobial therapy in the 1940s (9). Convalescent plasma was used during the 1918 flu epidemic and reduced mortality among plasma recipients (10). More recently, two other epidemics caused by coronaviruses have been associated with high mortality, severe acute respiratory syndrome coronavirus 1 (SARS-CoV-1) in 2003 and Middle East respiratory syndrome (MERS) in 2012. The SARS-CoV-1 epidemic was contained, but MERS became endemic in the Middle East and triggered a secondary major outbreak in South Korea. In both viral outbreaks, the high mortality and absence of effective therapies led to the use of convalescent plasma. In the largest study of the SARS-CoV-1 outbreak, among 80 patients in Hong Kong (11), patients treated within the first 14 days of infection had earlier discharge from hospital. These results are consistent with the notion that convalescent plasma may be an effective treatment of coronavirus infections and that earlier administration is more likely to be successful.

Although promising, convalescent plasma has not yet been demonstrated to be safe as a treatment for COVID-19. Thus, we analyzed key safety metrics following transfusion of convalescent plasma in 5,000 hospitalized adults with severe or life-threatening COVID-19. We hypothesized that the rate of serious adverse events related to the transfusion of convalescent plasma *per se* would be low and that the seven-day mortality rate would not be demonstrably elevated compared to other experiences with this deadly disease.

## Results

### EAP Participation

From April 3 to May 11, 2020, a total of 14,288 patients with severe or life-threatening COVID-19 or who were judged by a healthcare provider to be at high risk of progression to severe or life-threatening COVID-19 were enrolled in the EAP. In that time, a total of 8,932 enrolled patients received a COVID-19 convalescent plasma transfusion, **Figure 1**. Data from the first 5,000 transfused patients were included in this report.

**Figure 1.**
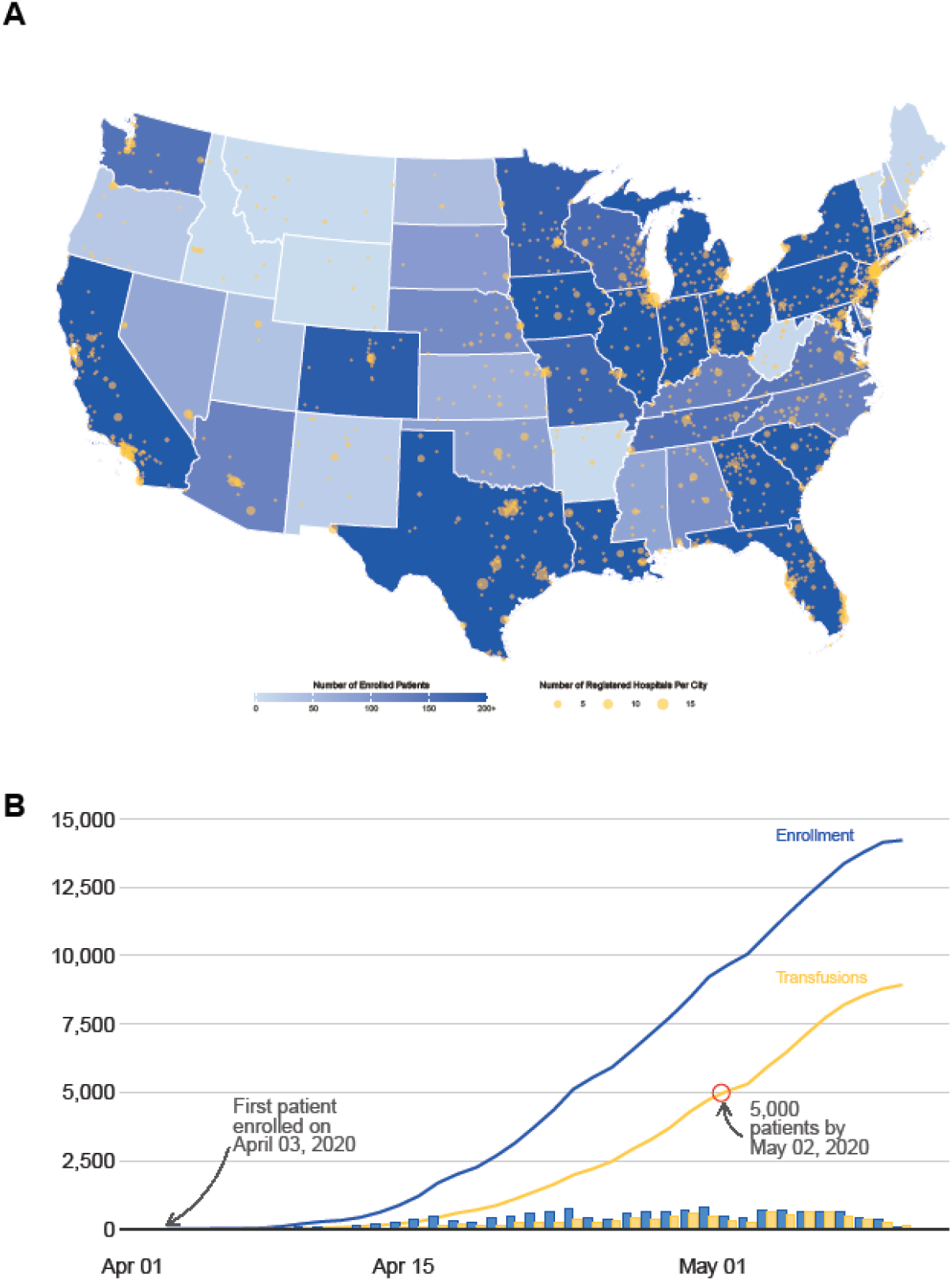
Participation in the US COVID-19 Convalescent Plasma Expanded Access Program (EAP) including data extracted on May 11, 2020. **A**. Choropleth map displaying the number of cumulatively enrolled patients in the EAP within each state of the contiguous US, with lower enrollment values displayed in a lighter hue and higher enrollment values displayed in a darker hue of blue. Registered acute care facilities are represented as filled yellow circles, with larger circles indicating greater number of registered facilities within the metropolitan area of a city. The choropleth map does not display data from non-contiguous US locations, including registered facilities in Puerto Rico, Hawaii, Alaska, Guam, and Northern Mariana Islands. **B**. The chronological line charts represent the cumulative number of enrolled patients (blue line) and the cumulative number of patients that have received a COVID-19 convalescent plasma transfusion (yellow line). The chronological bar charts represent analogous values— the number of enrolled patients (blue bars) and number of patients that have received a COVID 19 convalescent plasma transfusion (yellow bars) by day. The difference between the blue and yellow bars highlights a fulfillment gap in COVID-19 convalescent plasma, which was most acute at the onset of the EAP and has substantially improved.

### Demographics

Key demographic characteristics of the patients are presented in **Table 1**. The data set included 3,153 men, 1,824 women and 23 persons in other gender/sex categories with diverse racial representation including Asian (6%), American Indian or Alaskan Native (<1%), Black (18%), White (49%), Native Hawaiian or Pacific Islander (<1%) and Multi-racial (<1%). The median age was 62 years (range, 18 – 97 years).

**Table 1.**
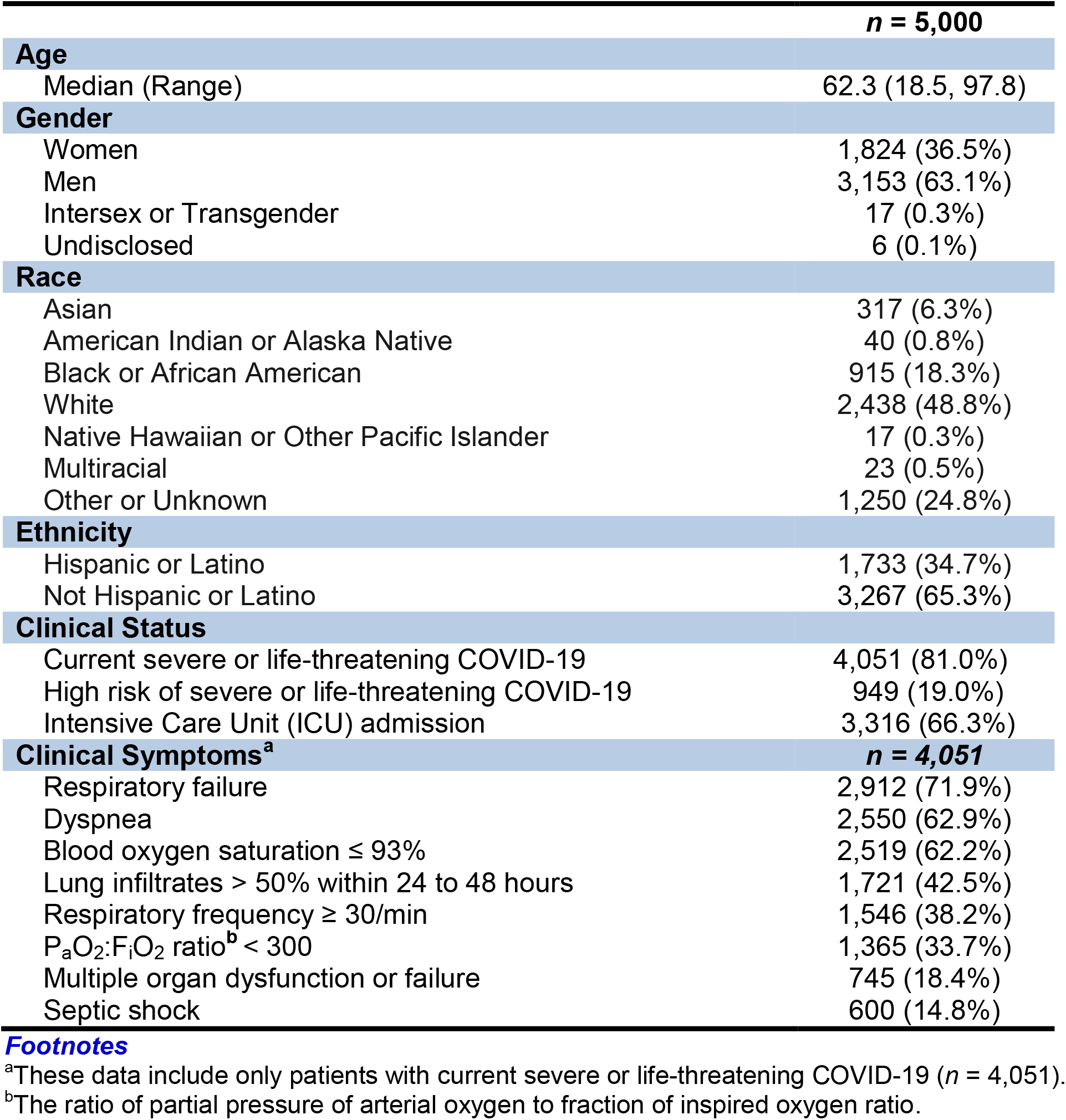
Patient Characteristics.

### Clinical Status and Symptoms

At the time of enrollment, 4,051 (81%) patients had severe or life-threatening COVID-19 and 949 (19%) were judged to have a high risk of progressing to severe or life-threatening COVID-19. Prior to COVID-19 convalescent plasma transfusion, a total of 3,316 patients (66%) were admitted to the ICU. Of the 4,051 patients diagnosed with severe or life-threatening COVID-19, 72% had respiratory failure, 63% reported dyspnea, 62% had a blood oxygen saturation ≤ 93%, 43% had lung infiltrates >50% within 24-28 hours of enrollment, 38% had a respiratory frequency ≥ 30 breaths·minute^−1^, 34% had partial pressure of arterial oxygen to fraction of inspired oxygen ratio < 300, 18% had multiple organ dysfunction or failure, and 15% had septic shock.

### Serious Adverse Events

Within four hours of completion of the COVID-19 convalescent plasma transfusion (inclusive of the plasma transfusion), 36 serious adverse events (SAEs) were reported (<1% of all transfusions). The attribution scale used by the treating physicians for evaluating the SAEs included unrelated, possibility related, probably related, or definitely related. Of the SAEs, 15 deaths were reported (0.3% of all transfusions) and four of those deaths were judged as related (possibly, *n*=3; probably, *n*=1; definitely, *n*=0) to the transfusion of COVID-19 convalescent plasma. There were 21 non-death SAEs reported, with seven reports of transfusion-associated circulatory overload (TACO), eleven reports of transfusion-related acute lung injury (TRALI), and three reports of severe allergic transfusion reaction. All incidences of TACO and TRALI were judged as related (possibly, n=9; probably, n=7; definitely, n=2) to the transfusion of COVID-19 convalescent plasma. The SAEs and their attributions are summarized in **Table 2**.

**Table 2.** Serious Adverse Event (SAE) Characteristics. (n=5,000)

| **Four Hour Reports** | **Reported (n = 36)** | **Related<sup>a</sup> (n = 25)** | **Estimate (95% CI)** |
| --- | --- | --- | --- |
| Mortality | 15 | 4 | 0.08% (0.03%, 0.21%) |
| Transfusion-Associated Circulatory Overload (TACO) | 7 | 7 | 0.14% (0.07%, 0.29%) |
| Transfusion-Related Acute Lung Injury (TRALI) | 11 | 11 | 0.22% (0.12%, 0.39%) |
| Severe allergic transfusion reaction | 3 | 3 | 0.06% (0.02%, 0.18%) |
| **Seven Day Reports** | **Reported** |  | **Estimate (95% CI)<sup>b</sup>** |
| Mortality | 602 |  | 14.9% (13.8%, 16.0%) |

<sup>a</sup>This category of serious adverse events (SAE) reports the aggregate total of possibly-, probably- and definitely-related SAEs, as attributed based on the site investigator's determination. The estimate is based on the number of related SAEs relative to the denominator of 5,000.
<sup>b</sup>The estimated seven-day mortality rate is based on a Kaplan-Meier estimate using all reported deaths. See methods for further estimation details including handling of censoring due to ongoing data collection.

Over the first seven days after the convalescent plasma transfusion, a total of 602 mortalities were observed. The overall seven-day mortality rate was estimated to be 14.9% (95% CI: 13.8%, 16.0%) using the product limit estimator; an estimate that was numerically higher than the crude estimate of 12.0% at day 7. Of the 3,316 patients admitted to the ICU, 456 mortalities were observed (16.7%, 95% CI: 15.3%, 18.1%). Of the 1,682 hospitalized patients not-admitted to the ICU, 146 mortalities were observed (11.2%, 95% CI: 9.5%, 12.9%).

## Discussion

### Safety Summary

In this initial report of 5,000 hospitalized patients in the US with severe or life-threatening COVID-19, or who were judged by a healthcare provider to be at high risk of progressing to severe or life-threatening COVID-19, the overall frequency of SAEs within four hours following the transfusion of COVID-19 convalescent plasma was less than 1% (*n* = 36) and the seven-day mortality rate was 14.9%. Although 70% of these SAEs were deemed to be related to plasma transfusion by treating physicians, most of the SAEs (56%) were judged as *possibly* related, suggesting uncertainty about the role of the transfusion *per se* in the adverse reaction. Additionally, the rate of SAEs definitely related to transfusion was objectively low (*n* = 2, <0.1% of all transfusions).

Although this study was not designed to evaluate efficacy of convalescent plasma we note with optimism the relatively low mortality in treated patients. The case fatality rate of COVID-19 has been reported to be ~4% among all persons diagnosed with COVID-19 (2); however, the case fatality rate among hospitalized patients is much higher ~15-20% (3, 5) and even more so among patients admitted to the ICU (57%) (4). Thus, the seven-day mortality rate was 14.9% reported here is not alarming, particularly because some of these plasma transfusions may be characterized as attempts at rescue or salvage therapy in patients admitted to the ICU with multi-organ failure, sepsis and significant comorbidities.

Despite these early and encouraging safety signals, there are several risks of COVID-19 convalescent plasma transfusion in critically-ill patients that warrant attention in this initial assessment of safety (12, 13).

### Transfusion-related acute lung injury (TRALI) and transfusion-associated circulatory overload (TACO)

The highest risk of mortality following plasma transfusion is likely due to sequelae pulmonary complications (14), and this risk is probably exacerbated by the underlying respiratory distress associated with COVID-19. Transfusion-related acute lung injury (TRALI) and transfusion-associated circulatory overload (TACO) are the two leading causes of transfusion-related mortality, and are often difficult to distinguish. These conditions have been emphasized in the plasma transfusion literature, but making an unequivocal determination of plasma-related toxicity in critically ill individuals is difficult in the face of ongoing conditions that resemble transfusion SAEs. Consequently, it is likely that some of the reported SAEs represent natural progression of the ongoing pathological processes.

The most common adverse event associated with plasma transfusion in critically-ill patients is TACO, which results in pulmonary edema and left atrial hypertension subsequent to circulatory overload. The reported incidence of TACO includes a large range from 1 in 14,000 in surveillance surveys to 12% in prospective studies in higher risk populations, showing the dependence of incidence on the clinical status of the transfusion recipient (15-17). TRALI often presents as bilateral pulmonary edema with little evidence of circulatory overload, and TRALI is further categorized into two types based on the absence of acute respiratory distress syndrome (ARDS) risk factors (type I) or presence of ARDS risk factors (type II) (18). The reported incidence of TRALI similarly covers a large range from ~0.01% in surveillance surveys to 8% in prospective studies of the critically ill (19, 20). The underlying lung injury associated with COVID-19 further complicates the differential diagnosis of TACO and TRALI, and may exacerbate the risk of transfusion-related reactions in these critically-ill patients. Although the incidence of transfusion-related reactions (TACO and TRALI) among critically-ill patients may be anticipated to be nearly 10%, the current data demonstrate an overall rate of reported transfusion-related serious adverse events less than 1%. Thus, the low rates of TRALI and TACO along with the "possibly related” attribution of most cases are reassuring.

### Antibody-Dependent Enhancement (ADE)

A theoretical concern of the use of COVID-19 convalescent plasma in patients with COVID-19 is a deteriorated clinical condition after plasma transfusion secondary to antibody-dependent enhancement (ADE) of infection or antibody-mediated proinflammatory effects (21). This theoretical concern is supported by reports of ADE in macaques given specific antibody administration prior to SARS-CoV-1 experimental infection (22) and ADE effects with other coronaviruses (23, 24). There is also the concern that antibody administration to individuals with significant viral loads may lead to the formation of antigen-antibody immune complexes, which may contribute to proinflammatory immune responses (25, 26). Although the specific signs and symptoms of ADE in humans with coronavirus infection are unknown, such an effect would presumably be associated with clinical deterioration and/or worse outcomes following convalescent plasma administration. The absence of a toxicity signature with the use of convalescent plasma in individuals with COVID-19 implies that this phenomenon may be clinically inconsequential. COVID-19 is known to elicit high neutralizing antibody titers in individuals who have recently recovered from infection and three case series of convalescent plasma administration also describe no deleterious ADE effects after infusion (27-29). The absence of untoward antibody-related effects after convalescent plasma administration could be due to the preferential binding of the neutralizing antibody to the virus rather than to immune cells or tissues which would be needed to enhance the proinflammatory immune responses responsible for ADE (30). Despite the absence of an apparent toxic effect attributable to specific antibody administration thus far, we caution continued vigilance as the use of antibody-based therapies and the number of treated individuals expands, particularly because specific high-risk groups may emerge that were not discernable in this initial cohort.

### Transfusion reactions and coagulation derangements

Another theoretical risk for convalescent plasma use in COVID-19 is the possibility that it could exacerbate the type of coagulation derangements associated with advanced COVID-19 (31). Absence of clinical outcomes related to severe thrombotic events within the four-hour SAE reports suggests that administration of 1-2 units of convalescent plasma does not *acutely* exacerbate potentially underlying disordered coagulation among critically-ill COVID-19 patients.

### Limitations

A key limitation of our observations includes the lack of detailed training of study personnel and monitoring in a highly diverse group of sites ranging from small community hospitals in rural areas to urban public hospitals to full-service academic medical centers. Given the speed at which the EAP was implemented and considering the stress on clinical staff at participating sites during this on-going pandemic, the web-based case reporting forms were designed to optimize convenience. Additionally, although the patient inclusion criteria were specific to hospitalized patients, these criteria were exceptionally broad. While these elements of the EAP may be suboptimal, they are perhaps understandable in a crisis of the magnitude of the COVID-19 pandemic.

The efficacy of convalescent plasma for treatment of COVID-19 has not yet been determined, and this report, focused on safety signals, should not be misconstrued as evidence of effectiveness. To test the efficacy of this therapy, future analyses of EAP data will include exposure control cohorts of patients who did not receive COVID-19 convalescent plasma. However, randomized controlled trials— some of which are currently in progress— will ultimately be necessary to evaluate the potential efficacy of convalescent plasma treatment along the continuum of disease-severity (http://ccpp19.org). Importantly, evolving data from the EAP will continue to have high utility in understanding the real world safety of COVID-19 convalescent plasma.

### Conclusion

In summary, the experience from the first 5000 patients with COVID-19 transfused with convalescent plasma provides no signal of toxicity beyond what is expected from plasma use in severely ill patients. Additionally, given the deadly nature of COVID-19 and the large population of critically-ill patients with multiple comorbidities included in these analyses, the mortality rate does not appear excessive. We also note that the data were reviewed by an independent Data and Safety Monitoring Board and have been deposited with the FDA and at no time was there consideration of stopping this therapy. Given the accelerating deployment of this therapy, these emerging data provide early safety indicators of convalescent plasma for COVID-19 treatment and suggest research should shift focus toward determining the efficacy of convalescent plasma.

## Methods

### Design and Oversight

The program is an FDA-initiated, national, multicenter, open-label Expanded Access Program (EAP) in hospitalized adults with severe or life-threatening COVID-19, or who were judged by a healthcare provider to be at high risk of progression to severe or life-threatening COVID-19. Initial discussions between the FDA and the Mayo Clinic related to the EAP began on March 30^th^, 2020. The program was approved by the Mayo Clinic Institutional Review Board (IRB) on April 1^st^, 2020 which served as the central IRB for all participating facilities and empaneled an independent Data and Safety Monitoring Board to oversee the safety analysis. All hospitals or acute care facilities in the US (including territories) were eligible to participate. Any willing, licensed US physician could participate as a treating physician-Principal investigator, provided they agreed to adhere to the treatment protocol, the terms of the FDA 1572 form, and all appropriate federal and state regulations. Registration occurred through the EAP central website, www.uscovidplasma.org.

The administrative and compliance infrastructure to implement the EAP was rapidly developed, and the initial web-based registration, compliance and data-entry system went live on April 3^rd^, 2020. The first patient received convalescent plasma on April 7^th^, and more than 5,000 hospitalized patients with COVID-19 were transfused with convalescent plasma under the EAP by May 3^rd^. **Figure 1** illustrates that over 2,000 acute care facilities have been registered and over 10,000 patients have been enrolled in the EAP across all 50 states and multiple US territories. Written informed consent was obtained from the participant or a legally-authorized representative prior to enrollment, except in jurisdictions allowing deferral of consent for emergency treatment, in which case, consent was obtained to continue participation.

### Participants

Eligible patients were aged 18 years or older, hospitalized with a laboratory confirmed diagnosis of infection with severe acute respiratory syndrome coronavirus 2 (SARS-CoV-2), and had, or were judged by a healthcare provider to be at high risk of progression to, severe or life-threatening COVID-19. Severe or life-threatening COVID-19 is defined by one or more of the following criterion: dyspnea, respiratory frequency ≥ 30 breaths·minute^-1^, blood oxygen saturation ≤ 93%, lung infiltrates >50% within 24-28 hours of enrollment, respiratory failure, septic shock, and multiple organ dysfunction or failure.

### Procedures

ABO-compatible COVID-19 convalescent plasma (200 – 500 mL) was administered intravenously according to institutional transfusion guidelines. Patients were continuously monitored with clinical assessments. Web-based standard data reporting surveys were completed 4-hours and 7-days post-transfusion, with additional forms used to report serious adverse events using the Research Electronic Data Capture (REDCap) system. All serious adverse event reports will be independently adjudicated over the course of the study by the IND Sponsor and trained designee (AMK) using the National Healthcare Safety Network Biovigilance Component Hemovigilance Module Surveillance Protocol as a framework (12).

### Outcomes

The primary outcome was to determine the safety of transfusion of COVID-19 convalescent plasma assessed as the incidence and relatedness of serious adverse events including death.

### Statistics

To facilitate the rapid enrollment of participants, sites and investigators, an electronic data collection system hosted at Mayo Clinic was built using the Research Electronic Data Capture System (REDCap, v.9.1.15 Vanderbilt University, Nashville, TN) (32, 33). Raw data were retrieved from REDCap via the application programming interface (API) and subjected to data consistency checks. Data presented in this initial safety report may undergo additional data quality control measures as the study progresses. The proportion of people that experienced one of a series of previously defined serious adverse events (SAEs) was summarized using a point estimate and 95% score confidence interval. To assess mortality, the time (in days) between transfusion and death was examined using the Kaplan Meier product limit estimator. Participants were censored at their last known vital status and all reported deaths through seven days were used to estimate the survival function. Data were censored at 0.25 days for patients that did not have follow-up beyond the initial report at four hours post transfusion at time of the analysis. For patients that expired within 24 hours, a survival time of 0.5 days was assigned. Precise time of day for key events was not recorded in the data collection system; thus, these imprecise time estimates were used. The point estimate and 95% CI were estimated at day 7 based on the estimated survival function. All analyses and graphics were produced with R version 3.6.2 (Vienna, Austria).

## Data Availability

Data are not publicly available at this time.

https://uscovidplasma.org

## Author Contributions

MJJ, NP, AC conceptualized the study in collaboration with the US FDA. MJJ, KAB, DF, ERW, AJC developed data reporting tools. REC, AMK, RFR, MNV, JEB reviewed and evaluated SAE reports. SEB, KFR, JGR, KJA, PRB, CMVB, JLW, JRS conceptualized data reporting metrics. REC, DOH, MRS, ERL, PWJ, MRB, KLK, JJD, RJR analyzed the data. MJJ, REC, JWS, SAK, RSW, AMK interpreted the results. MJJ, JWS, SAK, DF, AMK, CCW, JRAS, NP, AC wrote the manuscript. All authors reviewed and edited the manuscript. RSW oversaw, developed and coordinated the IRB and compliance related infrastructure required to rapidly initiate the EAP in parallel with the activities above.

## Acknowledgements

This study was supported in part by a US Department of Health and Human Services (HHS), Biomedical Advanced Research and Development Authority (BARDA) grant 75A50120C00096 (to MJJ), National Center for Advancing Translational Sciences (NCATS) grant UL1TR002377, National Heart, Lung, and Blood Institute (NHLBI) grant 5R35HL139854 (to MJJ), National Institute of Diabetes and Digestive and Kidney Diseases (NIDDK) 5T32DK07352 (to JWS and CCW), Natural Sciences and Engineering Research Council of Canada (NSERC) PDF-532926-2019 (to SAK), National Institute of Allergy and Infectious Disease (NIAID) grants R21 AI145356 and R21 AI152318 (to DF), R01 AI152078 9 (to AC), National Heart Lung and Blood Institute RO1 HL059842 (to AC), Schwab Charitable Fund (Eric E Schmidt, Wendy Schmidt donors), United Health Group, National Basketball Association (NBA), Millennium Pharmaceuticals, Octopharma USA, Inc, and the Mayo Clinic.

We thank the members of the Mayo Clinic Institutional Review Board, the Mayo Clinic Office of Human Research Protection, the Mayo Clinic Office of Research Regulatory support and in particular Mark Wentworth, the Executive Dean of Research at Mayo Clinic Dr. Gregory Gores and the CEO of Mayo Clinic Dr. Gianrico Farrugia for their support and assistance, and the independent Data and Safety Monitoring Board for their work and oversight of the Expanded Access Program— Dr. Allan S. Jaffe (chair), Dr. David O. Warner, Dr. William G. Morice II, Dr. Paula J. Santrach, Dr. Robert L. Frye, Dr. Lawrence J Appel, Dr. Taimur Sher. We thank the members of the National COVID-19 Convalescent Plasma Project (http://ccpp19.org) for their intellectual contributions and support. We thank the participating medical centers and medical teams, and blood centers for their rigorous efforts necessary to make this program possible. We also thank the donors for providing COVID-19 convalescent plasma.

